# Neutrophil percentages in bronchoalveolar lavage fluid: Implications for diagnosing bacterial pneumonia in patients with immunocompromise and neutropenia

**DOI:** 10.1101/2024.05.04.24306709

**Authors:** Kevin M. Grudzinski, Samuel Fenske, Alec Peltekian, Nikolay S. Markov, Anna Pawlowski, Mengjia Kang, James M. Walter, Chiagozie I. Pickens, Nandita R. Nadig, Ankit Agrawal, Benjamin D. Singer, Richard G. Wunderink, Catherine A. Gao, NU SCRIPT Study Investigators

**Author notes:** Address correspondence to: Catherine A. Gao, MD MS, Assistant Professor of Medicine, Division of Pulmonary and Critical Care, Northwestern University Feinberg School of Medicine, 625 N Michigan Ave, 15^th^ floor, Chicago, Illinois 60611. **Human Ethics and Consent to Participate:** This study was approved by the Northwestern University Institutional Review Board with study ID STU00204868. Study participants or their surrogates provided informed consent. **Data availability:** A significant portion of this data has been already made available through PhysioNet at https://physionet.org/content/script-carpediem-dataset/1.1.0/, a future update will include new patients and updated data since the publication of the original dataset. Code associated with this project is available at https://github.com/NUSCRIPT/grudzinski_neutropenia_2024.

## Abstract

**Rationale:** Pneumonia is the most common infection in ICU patients and a leading cause for death. Assessment of bronchoalveolar lavage fluid (BALF) cellularity can aid in pneumonia diagnosis. Low percentages (<50%) of BALF neutrophils have a high negative predictive value for bacterial pneumonia in a general medical ICU population. The operating characteristics in patients with immunocompromise and neutropenia are less clear.

**Objective:** To compare BALF % neutrophils operating characteristics in patients with and without immunocompromise or neutropenia.

**Methods:** This was a single center observational cohort study. Patients were categorized into three groups: (1) patients with neutropenia, (2) patients with underlying immunocompromise, and (3) patients with neither. BAL-level analysis reflected neutropenia and immunocompromise state on day of BAL sampling. Operating characteristics of BALF % neutrophils were calculated using varying thresholds of alveolar neutrophilia. Median [Q1,Q3] are reported for nonparametric data and compared using Mann-Whitney U tests.

**Results:** 688 mechanically ventilated patients had 1736 BAL samples. Among bacterial pneumonia episodes, no difference was found in BALF % neutrophils between patients with underlying immunocompromise and patients with neither neutropenia nor immunocompromise on day of sampling: 82.0% [61.0, 91.0] vs 81.0% [66.0, 91.0], p=0.859. However, when neutropenic on day of sampling, the median BALF % neutrophils was 35.0% [8.8, 67.5] (p<0.001 compared with other categories). In patients with neutropenia, a BALF % neutrophil threshold of 7% had a sensitivity of 90% for excluding bacterial pneumonia.

**Conclusions:** We found that among patients with bacterial pneumonia, BALF % neutrophil was not significantly lower in patients with a broad spectrum of immunocompromised states but was significantly lower when measured during acute neutropenia. We found varying thresholds of BALF % neutrophils across the three groups. Patients with neutropenia who mount even a low percent of alveolar neutrophils should raise concern for bacterial pneumonia.

## Introduction

Pneumonia is the leading cause of deaths from infection and the most common infection identified in intensive care unit (ICU) patients. Severe pneumonia requiring ICU admission is associated with high rates of morbidity and mortality.^1,2^ Detection of pneumonia in critically ill patients is complex, and uncertainty in diagnosing the presence and cause of pneumonia often leads to the unnecessary administration of antibiotic therapy to uninfected patients.^3,4^ Assessment of bronchoalveolar lavage fluid (BALF) cellularity can aid in pneumonia diagnosis. Recruitment of neutrophils into the alveolus is a critical component of host defense to bacterial infection. Low percentages (<50%) of BALF neutrophils have been previously shown to have a high negative predictive value for bacterial pneumonia in a general medical ICU population.^4,5^ How to interpret BALF % neutrophils in immunocompromised patients, especially neutropenic patients, is less clear.

Patients can have a variety of immune defense disorders, encompassing innate, cellular, and humoral immune mechanisms. These immunocompromising conditions, including neutropenia, put the host at increased risk of infection.^6^ Clinically, immunocompromised and neutropenic hosts are often risk stratified as having unique infectious risks due to differences in the underlying causes of their immunocompromise and the specific pathways that are involved.^7,8^ Classical clinical signs of pneumonia, including new pulmonary infiltrates, leukocytosis, fever, and purulent secretions may be less apparent in immunocompromised patients, adding to the challenge of appropriately identifying patients who are likely to benefit from further investigation and empiric antimicrobial therapy.^9^ However, these patients can develop multiple non-infectious conditions that can mimic pneumonia including aspiration pneumonitis, pulmonary edema, drug toxicity, graft-versus host disease, and others.^10^ Neutropenia adds to the differential diagnosis of pulmonary infiltrates, which may arise from both infectious and non-infectious causes such as leukemic infiltrate, drug toxicity, or systemic response to disease-directed therapy, though these considerations are not exclusive to neutropenic patients.^9^

Non-invasive testing does not always yield a definitive diagnosis in mechanically ventilated patients with suspected pneumonia, ^11^ highlighting the importance of obtaining BALF for definitive diagnosis. The operating characteristics of BALF % neutrophils in patients with immunocompromise and neutropenia have been previously examined in small cohorts^12^ but not tied to pneumonia diagnosed by state-of-the-art BALF microbiology, including multiplex PCR, with a standardized multi-physician review process. Thus, our aim was to compare BALF % neutrophils operating characteristics in cases of pneumonia, focusing on discerning differences between patients with neutropenia, immunocompromise, or neither.

## Patients and Methods

### Patient Cohort

We analyzed BALF specimens obtained as part of the clinical evaluation of known or suspected pneumonia in mechanically ventilated patients to discern differences between individuals with and without immunocompromise or neutropenia. The cohort was enrolled in the single-center Successful Clinical Response in Pneumonia Therapy (SCRIPT) study. Patients’ families or legally authorized representative consent to partake in this observational cohort study.

### Pneumonia Episode Adjudication

At our institution, bronchoalveolar lavage (BAL) or non-bronchoscopic BAL (NBBAL) is routinely and safely performed to confirm presence and etiology of pneumonia in intubated patients.^13^ NBBALs are performed as a respiratory therapist-driven protocol.^13^ BALF neutrophils are reported as a percentage of leukocytes by the clinical laboratory. Fungal pneumonia episodes were not included in our cohort. Episode-defining BALs are the first BAL associated with a new pneumonia episode. Patients often had multiple BALs collected during their course and not all were episode-defining, as they may have been collected during an ongoing episode. The timing and etiology of each pneumonia episode (linked to an episode-defining BAL, see Figure 1A for an illustrative hypothetical case) was adjudicated by a committee of critical care physicians using a predefined protocol, with episode etiologies including bacterial, bacterial/viral, viral or non-pneumonia.^13,14^ A patient’s BAL level data could also span different categories based on blood neutrophil count at time of sampling during a particular pneumonia episode. Immunocompromised state (such as hematologic malignancy, organ transplant, receiving chronic immunosuppressing medications, etc., see Supplemental Material 1 for full list) was categorized by the research intake team reviewing the chart and interviewing family members on study intake as described in previous work. ^15^ Neutropenia was defined as a peripheral absolute neutrophil count (ANC) <1500 cells/µL. Patients were categorized into three groups based on immunocompromised state at study entry and whether they experienced neutropenia during their ICU admission with the following order of priority: patients with neutropenia, patients with underlying immunocompromise, and patients with neither. BAL-level analysis reflects neutropenia and immunocompromise state on day of BAL sampling.

**Figure 1.**
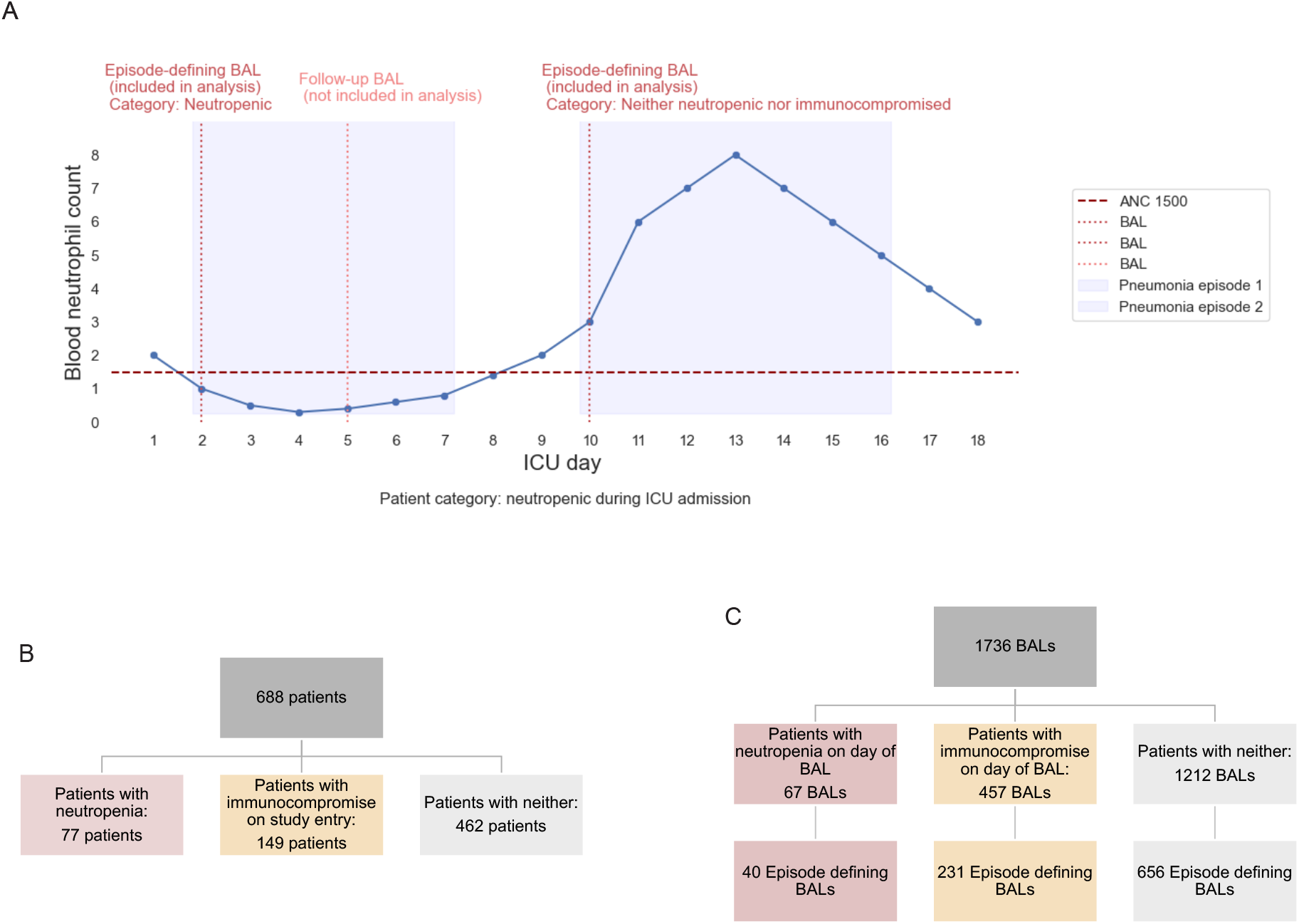
**(A) Patient course example**. An illustrative hypothetical course of a patient highlighting changes in their blood neutrophil counts over the ICU admission. A patient with neutropenia at any point during their ICU admission is categorized as ‘neutropenic’. For BAL level analysis, blood neutrophil count at the time of an episode-defining BAL, which is the BAL collected that the committee marked as the start of pneumonia episode, determined their categorization for analysis. A patient’s BAL data included in analysis could therefore span the different categories based on their blood neutrophil count at time of sampling and start of a new pneumonia episode. Follow-up BALs during a pneumonia episode were not included in the analysis. **(B) Overview of study design. Level one**: total patients. **Level two**: total number of patients split by whether they were neutropenic during admission, immunocompromised as defined by research team on study entry, or neither. **(C) Overview of BAL level data. Level one**: total number of BALs. **Level two:** total number of BALs obtained by group discerning if sample was collected on a day of neutropenia, collected from patients with underlying immunocompromise, or patients with neither. **Level three**: number of episode-defining samples by group

### Statistical Analysis

Data were abstracted from the electronic health record and study databases as previously described.^16^ Data are expressed as median (Quartile [Q] 1, Q3) and compared using Mann-Whitney U tests. Using episode-defining BALs and varying thresholds of BALF % neutrophils, we calculated sensitivity, specificity, negative predictive value (NPV), positive predictive value (PPV), positive likelihood ratio (+LR), and negative likelihood ratio (-LR) to identify the presence of bacterial pneumonia as outlined by multiplex PCR and culture results. Sensitivity and specificity were plotted at varying BALF % neutrophil thresholds for patients with neutropenia, patients with underlying immunocompromise, and patients with neither. Analysis done using Python v3.10.4. Code associated with this is available https://github.com/NUSCRIPT/grudzinski_neutropenia_2024.

## Results

### Demographic and Clinical Characteristics

Demographic and clinical characteristics of the 688 mechanically ventilated patients with known or suspected pneumonia included 409 (59.4%) males. Median age was 62 [51,71] (Table 1A). 77 patients (11.2%) experienced neutropenia during their ICU stay, 149 (21.7%) had underlying immunocompromise on study entry, and 462 (67.1%) had neither (Figure 1B). Common categories of immunocompromise were recent chemotherapy (43 patients, 6.2%), solid organ transplant (39 patients, 5.7%), stem cell transplant (36 patients, 5.2%), and acute leukemia (30 patients, 4.4%). The majority of patients experiencing neutropenia (n=63, 81.8%) had an underlying immunocompromising condition, but 14 (18.2%) patients did not have an underlying immunocompromising condition. These patients developed neutropenia during their admission with a short duration of neutropenic days (median 2 [1,4]). In 13 (92.9%) of these patients, their neutropenia was attributed to sepsis. Unfavorable outcomes of death, discharge to hospice, or lung transplantation during admission occurred in 49 (63.6%) patients with neutropenia, 83 (55.7%) patients with underlying immunocompromise, and 179 (38.7%) patients with neither.

**Table 1.** Characteristics of the study cohort. (**A)** Demographic and clinical characteristics of the study cohort. The ‘Immunocompromised’ category is inclusive of subcategories. Solid Organ Transplant, Stem Cell Transplant, Leukemia, and Chemotherapy, and were categorized by the research staff at the time of enrollment into the study, whereas neutropenia during admission was summarized separately based on bloodwork during ICU admission. Other categories of immunocompromise are available in Supplemental Material 1. Unfavorable outcomes are death, discharge to hospice, or requiring lung transplant. (**B)** Clinical characteristics of BAL samples, split by a patient’s first BAL during their ICU stay and subsequent BALs. Antibiotic and ventilator days are summed across ICU days prior to sampling. Not all subsequent BALs are episode-defining BALs.

|  | Patients with Neutropenia | Patients with Underlying Immunocompromise | Patients with Neither |
| --- | --- | --- | --- |
| **n** | 77 | 149 | 462 |
| **Age, median [Q1,Q3]** | 59.0 [48.0,69.0] | 63.0 [52.0,70.0] | 63.0 [51.0,72.0] |
| **Gender, male, n (%)** | 56 (72.7) | 77 (51.7) | 276 (59.7) |
| **Immunocompromised, n (%)** | 63 (81.8) | 149 (100.0) |  |
| Solid Organ Transplant, n (%) | 8 (10.4) | 31 (20.8) |  |
| Stem Cell Transplant, n (%) | 14 (18.2) | 22 (14.8) |  |
| Acute Leukemia, n (%) | 21 (27.3) | 9 (6.0) |  |
| Chemotherapy, n (%) | 24 (31.2) | 19 (12.8) |  |
| **WBC Count, median [Q1,Q3]** | 3.3 [1.4,7.7] | 11.5 [7.9,16.2] | 11.6 [9.1,14.8] |
| **Neutrophil Count, median [Q1,Q3]** | 2.3 [0.8,4.9] | 9.8 [6.8,14.4] | 9.3 [6.8,13.0] |
| **BAL % Neutrophils, median [Q1,Q3]** | 45.8 [10.2,72.8] | 72.5 [53.5,88.0] | 72.0 [51.0,85.1] |
| **Bacterial Episode, n (%)** | 49 (63.6) | 106 (71.1) | 342 (74.0) |
| **Cumulative ICU Days, median [Q1,Q3]** | 20.0 [13.0,34.0] | 14.0 [8.0,26.0] | 15.0 [8.0,28.0] |
| **Unfavorable Outcome, n (%)** | 49 (63.6) | 83 (55.7) | 179 (38.7) |

(B)
|  | **First BAL** | **Follow Up BALs** |
| --- | --- | --- |
| **BAL Count, n** | 688 | 1048 |
| **Day After First ICU Day, median [Q1,Q3]** | 1.0 [0.0,2.2] | 15.0 [8.0,29.2] |
| **Days On Ventilatory Before BAL, median [Q1,Q3]** | 1.0 [1.0,2.0] | 14.0 [8.0,25.0] |
| **Summed Days of Antibiotics, median [Q1,Q3]** | 2.0 [1.0,3.0] | 12.0 [7.0,21.0] |
| **Immune System Status on Day of Sampling, n (%)** |  |  |
| Patients with Neutropenia | 36 (5.2) | 31 (3.0) |
| Patients with Underlying Immunocompromise | 176 (25.6) | 281 (26.8) |
| Patients with Neither | 476 (69.2) | 736 (70.2) |
| **BALF Cell Breakdown** |  |  |
| BAL % Neutrophils, median [Q1,Q3] | 71.0 [41.0,88.0] | 77.0 [50.0,89.0] |
| BAL % Lymphocytes, median [Q1,Q3] | 5.0 [2.0,11.0] | 4.0 [2.0,9.0] |
| BAL % Macrophages, median [Q1,Q3] | 11.0 [4.0,28.0] | 10.0 [4.0,25.2] |
| **Episode Etiology, n (%)** |  |  |
| Bacterial | 218 (42.2) | 221 (53.8) |
| Bacterial/viral | 68 (13.2) | 130 (31.6) |
| Viral | 134 (26.0) | 42 (10.2) |
| Non-pneumonia control | 96 (18.6) | 18 (4.4) |
| **Episode Category, n (%)** |  |  |
| CAP | 144 (27.9) | 7 (1.7) |
| HAP | 196 (38.0) | 61 (14.8) |
| VAP | 80 (15.5) | 325 (79.1) |

### BAL Level Analysis

Features of the 1736 BAL samples performed are shown in Table 1B. Of 927 unique episodes of suspected pneumonia, 439 (47.4%) were adjudicated as bacterial, 198 (21.4%) combined bacterial/viral, 176 (18.9%) viral, and 114 (12.3%) were not consistent with pneumonia. Of these unique episodes, 151 (16.3%) were adjudicated as CAP, 257 (27.7%) HAP, and 405 (43.7%) VAP, defined at the time of BAL sampling. 67 BALs were obtained on patients during a neutropenic day with 40 episode-defining samples. 457 BALs were obtained from patients with underlying immunocompromise but not neutropenia on day of BAL, with 231 episode-defining samples. 1212 BALs were obtained from patients with neither neutropenia nor immunocompromise on day of BAL with 656 episode-defining samples (Figure 1C).

### Predictive value of BALF % Neutrophils

As expected, BALF % neutrophils correlated with peripheral neutrophil count on the day of sampling (Supplemental Figure 1).

Among pneumonia episodes classified as bacterial, no difference was found in BALF % neutrophils between patients with underlying immunocompromise and patients with neither on day of sampling: 82.0% [61.0, 91.0] vs 81.0% [66.0, 91.0], p = 0.859 (Figure 2A). However, when neutropenic on day of sampling, the median BALF % neutrophils was 35.0% [8.8, 67.5] (p<0.001 compared with the patients with underlying immunocompromise, p<0.001 compared with the patients with neither). Similarly, no difference was found among peripheral neutrophils between patients with underlying immunocompromise and patients with neither on day of sampling: 10.4 10^3^cells/µL [6.6, 15.9] vs 11.5 [7.3, 17.5], p = 0.128 (Figure 2B). However, when neutropenic, median peripheral neutrophils was 0.5 10^3^cells/µL [0.0, 0.7] (p<0.001 compared with the patients with underlying immunocompromise, p<0.001 compared with the patients with neither). Similar patterns were seen across viral pneumonia episodes. The combined viral/bacterial category had only three samples taken during neutropenic days.

**Figure 2.**
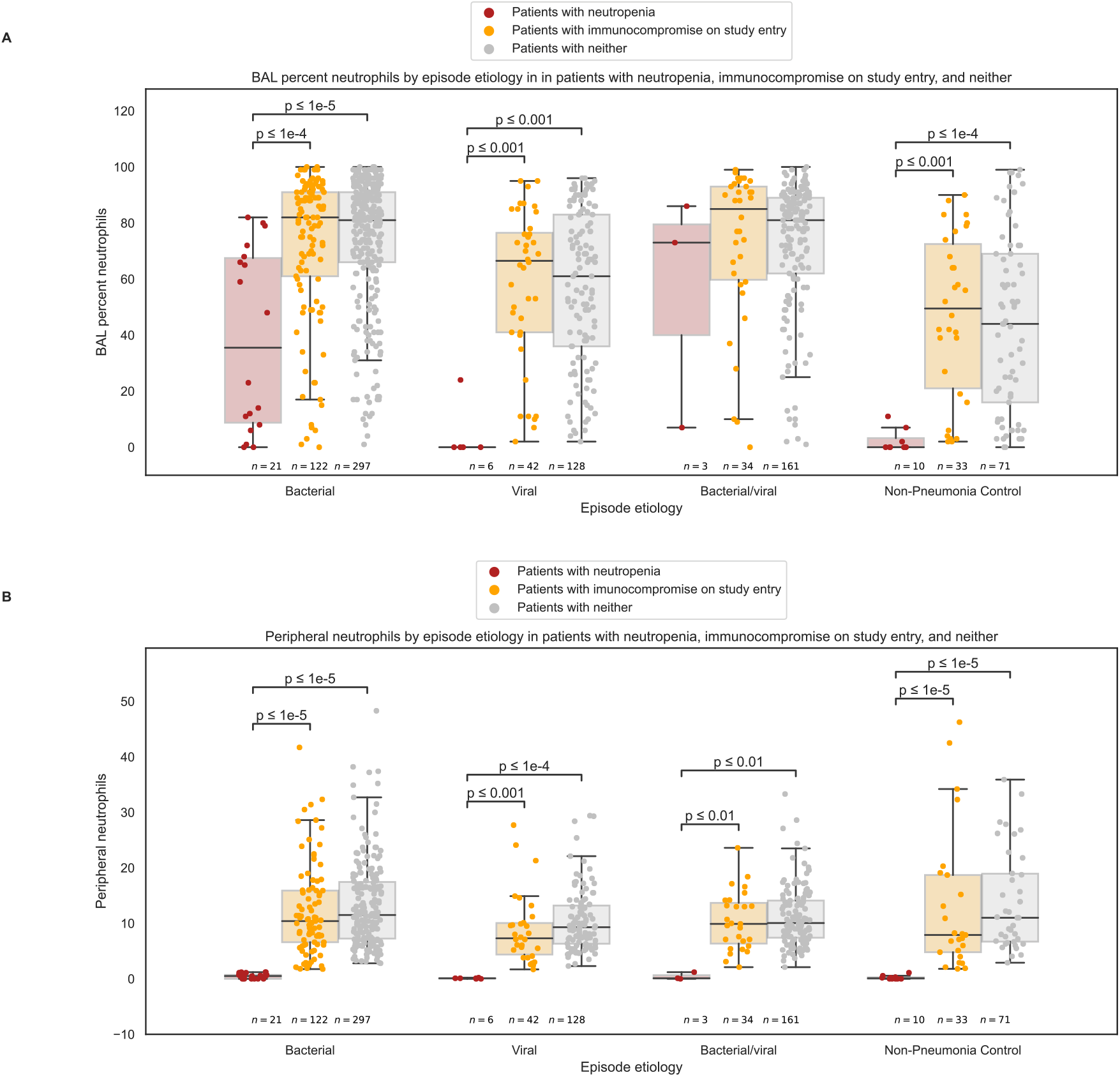
**(A)** Box plots showing BALF % neutrophils and interquartile range by pneumonia episode etiology. The data is split by patients with neutropenia, patients with underlying immunocompromise, or patients with neither on day of BAL sampling. Data were compared by pairwise Mann-Whitney U tests. **(B)** Box plots showing peripheral % neutrophils and interquartile range by pneumonia episode etiology on the same day as the BAL samples presented in Figure 1A. The data is split by patients with neutropenia, patients with underlying immunocompromise, or patients with neither based on their peripheral neutrophil count on day of BAL sampling. Data were compared by pairwise Mann-Whitney U tests.

Operating characteristics of BALF % neutrophils as a predictor of bacterial pneumonia across the three groups are shown in Table 2. In patients with neutropenia, the traditional BALF % neutrophil threshold of less than 50% had a sensitivity of 64.7%, specificity of 82.5%, PPV of 61.1% and NPV of 84.6%. For the patients with neutropenia, a BALF % neutrophil threshold of 7% had a sensitivity of 90% (Figure 3A). Whereas, for the patients with underlying immunocompromise and patients with neither, (Figure 3B & 3C) a BALF % neutrophil threshold of 29% and 35% had a sensitivity of 90%, respectively.

**Table 2.** Operating Characteristics of BALF % Neutrophils to Identify the Presence of Bacterial Pneumonia Across the Three Groups. (A) Operating Characteristics of BALF % Neutrophils to Identify the Presence of Bacterial Pneumonia in Patients with Neutropenia. (B) Operating Characteristics of BALF % Neutrophils to Identify the Presence of Bacterial Pneumonia in Patients with Underlying Immunocompromise. (C) Operating Characteristics of BALF % Neutrophils to Identify the Presence of Bacterial Pneumonia in Patients with Neither. *Definition of abbreviations: CI = confidence interval; LR = likelihood ratio; NPV = negative predictive value; PPV = positive predictive value.*

| Operating Characteristics of BALF % Neutrophils to Identify the Presence of Bacterial Pneumonia in Patients with Neutropenia |  |  |  |  |  |  |
| --- | --- | --- | --- | --- | --- | --- |
| BAL Neutrophils (%) | Sensitivity (%) | Specificity (%) | PPV (%) | NPV (%) | Positive LR (95% CI) | Negative LR (95% CI) |
| ≥10 | 75 | 57.7 | 35.3 | 88.2 | 1.77 (1.0 - 3.1) | 0.43 (0.20 - 0.93) |
| ≥20 | 75 | 73.1 | 46.2 | 90.5 | 2.79 (1.6 - 4.9) | 0.34 (0.17 - 0.71) |
| ≥30 | 75 | 80.8 | 54.5 | 91.3 | 3.9 (2.4 - 6.8) | 0.31 (0.15 - 0.64) |
| ≥40 | 75 | 80.8 | 54.5 | 91.3 | 3.9 (2.4 - 6.8) | 0.31 (0.15 - 0.64) |
| ≥50 | 75 | 84.6 | 60.0 | 91.7 | 4.88 (2.8 - 8.5) | 0.3 (0.15 - 0.60) |
| ≥60 | 75 | 88.5 | 66.7 | 92.0 | 6.5 (3.7 - 11.3) | 0.28 (0.140 - 0.57) |
| ≥70 | 62.3 | 96.2 | 83.3 | 89.3 | 16.3 (8.4 - 31.5) | 0.39 (0.19 - 0.78) |
| ≥80 | 25 | 96.2 | 66.7 | 80.6 | 6.5 (1.8 - 22.9) | 0.78 (0.39 - 1.57) |
| ≥90 | 0 | 100 | 0.0 | 76.5 | inf | 1 (0-0) |

(B)
| Operating Characteristics of BALF % Neutrophils to Identify the Presence of Bacterial Pneumonia in Patients with Underlying Immunocompromise |  |  |  |  |  |  |
| --- | --- | --- | --- | --- | --- | --- |
| BAL Neutrophils (%) | Sensitivity (%) | Specificity (%) | PPV (%) | NPV (%) | Positive LR (95% CI) | Negative LR (95% CI) |
| ≥10 | 95.1 | 9.6 | 48.8 | 68.8 | 1.05 (0.87 - 1.3) | 0.50 (0.28 - 0.91) |
| ≥20 | 94.2 | 17.5 | 50.8 | 76.9 | 1.14 (0.95 - 1.4) | 0.33 (0.21 - 0.52) |
| ≥30 | 90.3 | 19.3 | 50.3 | 68.8 | 1.12 (0.92 - 1.4) | 0.50 (0.33 - 0.77) |
| ≥40 | 88.3 | 22.8 | 50.8 | 68.4 | 1.15 (0.94 - 1.4) | 0.51 (0.35 - 0.75) |
| ≥50 | 82.5 | 32.5 | 52.5 | 67.2 | 1.22 (1.0 - 1.5) | 0.54 (0.39 - 0.75) |
| ≥60 | 75.7 | 40.4 | 53.4 | 64.8 | 1.27 (1.0 - 1.6) | 0.60 (0.45 - 0.81) |
| ≥70 | 68 | 53.5 | 65.9 | 64.9 | 1.46 (1.2 - 1.8) | 0.60 (0.46 - 0.78) |
| ≥80 | 56.3 | 67.5 | 61.1 | 63.1 | 1.74 (1.4 - 2.2) | 0.65 (0.51 - 0.82) |
| ≥90 | 34 | 85.1 | 67.3 | 58.8 | 2.28 (1.7 - 3.2) | 0.78 (0.63 - 0.96) |

(C)
| Operating Characteristics of BALF % Neutrophils to Identify the Presence of Bacterial Pneumonia in Patients with Neither |  |  |  |  |  |  |
| --- | --- | --- | --- | --- | --- | --- |
| BAL Neutrophils (%) | Sensitivity (%) | Specificity (%) | PPV (%) | NPV (%) | Positive LR (95% CI) | Negative LR (95% CI) |
| ≥10 | 97.6 | 7.4 | 55.6 | 72.4 | 1.05 (0.94 - 1.2) | 0.32 (0.21 - 0.49) |
| ≥20 | 94.4 | 13.7 | 56.5 | 67.2 | 1.09 (0.97 - 1.2) | 0.41 (0.30 - 0.56) |
| ≥30 | 93.2 | 20.4 | 58.1 | 71.6 | 1.17 (1.0 - 1.3) | 0.33 (0.26 - 0.43) |
| ≥40 | 89.3 | 27.1 | 59.3 | 68.1 | 1.23 (1.1 - 1.4) | 0.39 (0.32 - 0.49) |
| ≥50 | 83.1 | 33.5 | 59.7 | 62.5 | 1.25 (1.1 - 1.4) | 0.51 (0.42 - 0.62) |
| ≥60 | 78.3 | 43.7 | 62.3 | 62.9 | 1.39 (1.2 - 1.6) | 0.5 (0.42 - 0.59) |
| ≥70 | 70.3 | 55.6 | 65.3 | 61.2 | 1.59 (1.4 - 1.8) | 0.53 (0.46 - 0.62) |
| ≥80 | 55.8 | 69 | 68.1 | 56.8 | 1.80 (1.6 - 2.1) | 0.64 (0.56 - 0.73) |
| ≥90 | 29.4 | 85.2 | 68.8 | 50.1 | 1.85 (1.5 - 2.3) | 0.84 (0.75 - 0.94) |

**Figure 3.**
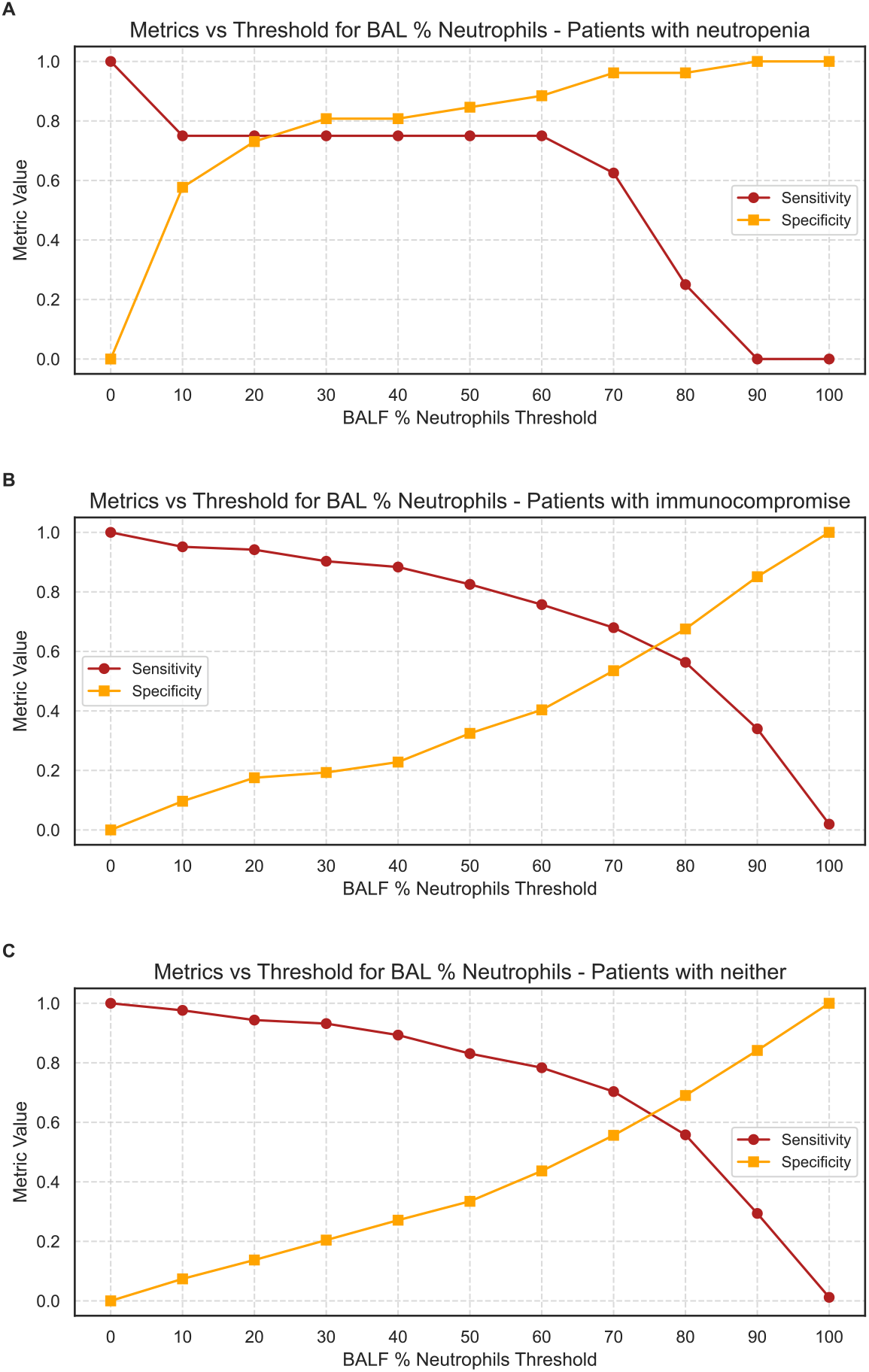
**(A)** Sensitivity and specificity plotted at varying BALF % neutrophils thresholds to identify the presence of bacterial pneumonia as outlined by multiplex PCR and culture results in patients with neutropenia **(B)** Sensitivity and specificity plotted at varying BALF % neutrophils thresholds to identify the presence of bacterial pneumonia as outlined by multiplex PCR and culture results in patients with underlying immunocompromise **(A)** Sensitivity and specificity plotted at varying BALF % neutrophils thresholds to identify the presence of bacterial pneumonia as outlined by multiplex PCR and culture results in patients with neither

## Discussion

We demonstrate an assessment of the host response in episodes of pneumonia in 688 patients who were neutropenic, underlying immunocompromise, or neither, utilizing nearly one thousand BAL samples. We show that among patients with bacterial pneumonia, BALF neutrophil percentage was not significantly lowered by a broad spectrum of immunocompromised states. The subset of patients who were neutropenic at the actual time of BAL sampling had significantly lower BALF % neutrophils.

Our study is the largest to determine the test characteristics of BALF % neutrophils in critically ill patients with neutropenia and immunocompromise.^4,17–19^ Assessment of BALF % neutrophils in patients with immunocompromise has similar sensitivity and specificity as previously reported that reported; therefore, it can help to rapidly exclude bacterial pneumonia in critically ill patients.^4^ Our study found that patients with neutropenia had a BALF % neutrophil threshold of 7% to achieve 90% sensitivity. Assessment of BALF % neutrophils in patients with neutropenia at time of sampling has a shift in the operating characteristics with the tradeoff of sensitivity and specificity occurring at a lower BALF % neutrophil threshold. Sensitivity is lower overall and begins declining at a lower threshold, indicating a greater diagnostic challenge in excluding bacterial pneumonia in this cohort. Therefore, patients with neutropenia who mount even a low percent of alveolar neutrophils should raise concern for bacterial pneumonia.

Overall, we found that patients with a wide range of immunocompromising conditions were able to mount an alveolar neutrophilic response in the setting of bacterial pneumonia. However, those with neutropenia mounted a less robust alveolar neutrophilia. Initial diagnosis of suspected pneumonia with rapid multiplex PCR or other biomarkers of host response may be more helpful in diagnosing infections in this population.

A small subset of patients experienced neutropenia without a clear underlying immunocompromising condition on study entry. These periods of neutropenia were often short in duration. Transient neutropenia, as may occur in the setting of severe sepsis or other critical illness, does not qualify patients as being considered inherently immunocompromised^6,20^ and therefore may not confer the same infectious risk as hosts with immunocompromising conditions.

Limitations to our study are that only mechanically ventilated patients were included and results may not apply to less ill patients. The subset of patients without typical underlying immunocompromised conditions who were neutropenic at the time of BAL only had a small number of episode-defining BALs, precluding further characterization of this population. In addition, we did not analyze the association between duration of neutropenia or prophylactic antibiotics, which are often administered to patients with prolonged neutropenia from hematological malignancies), prior to BAL sampling.

We propose a lower threshold of BALF % neutrophils to be used in patients who are neutropenic at time of sampling to suggest against bacterial pneumonia. Future work should focus on validating this threshold in a wider cohort of patients.

## Data Availability

A significant portion of this data has been already made available through PhysioNet at https://physionet.org/content/script-carpediem-dataset/1.1.0/, a future update will include new patients and updated data since the publication of the original dataset.

## Abbreviations List

(ICU): Intensive care unit
(BALF): bronchoalveolar lavage fluid
(NBBAL): non-bronchoscopic BAL
(ANC): absolute neutrophil count
(NPV): negative predictive value
(PPV): positive predictive value
(+LR): positive likelihood ratio
(-LR): negative likelihood ratio
(CAP): community associated pneumonia
(HAP): hospital associated pneumonia
(VAP): ventilator associated pneumonia

---

**Supplemental Material 1**: List of immunocompromised status/medications used by research team to flag patients as immunocompromised on study entry.

Medical Conditions:

Acute leukemia

HIV

Immunoglobulin deficiency

Lymphoma

Multiple myeloma

Solid organ transplant

Hematologic malignancy

Other malignancy

Non-malignancy

Medications:

Azathioprine

Chronic corticosteroids (last month) > 5 mg/d

Chronic corticosteroids (last month) >= 20 mg/d

Cyclosporine

Cyclophosphamide

Mycophenolate

Myelosuppressive chemotherapy

Rituximab

Tacrolimus

Other

**Supplemental Figure 1.**
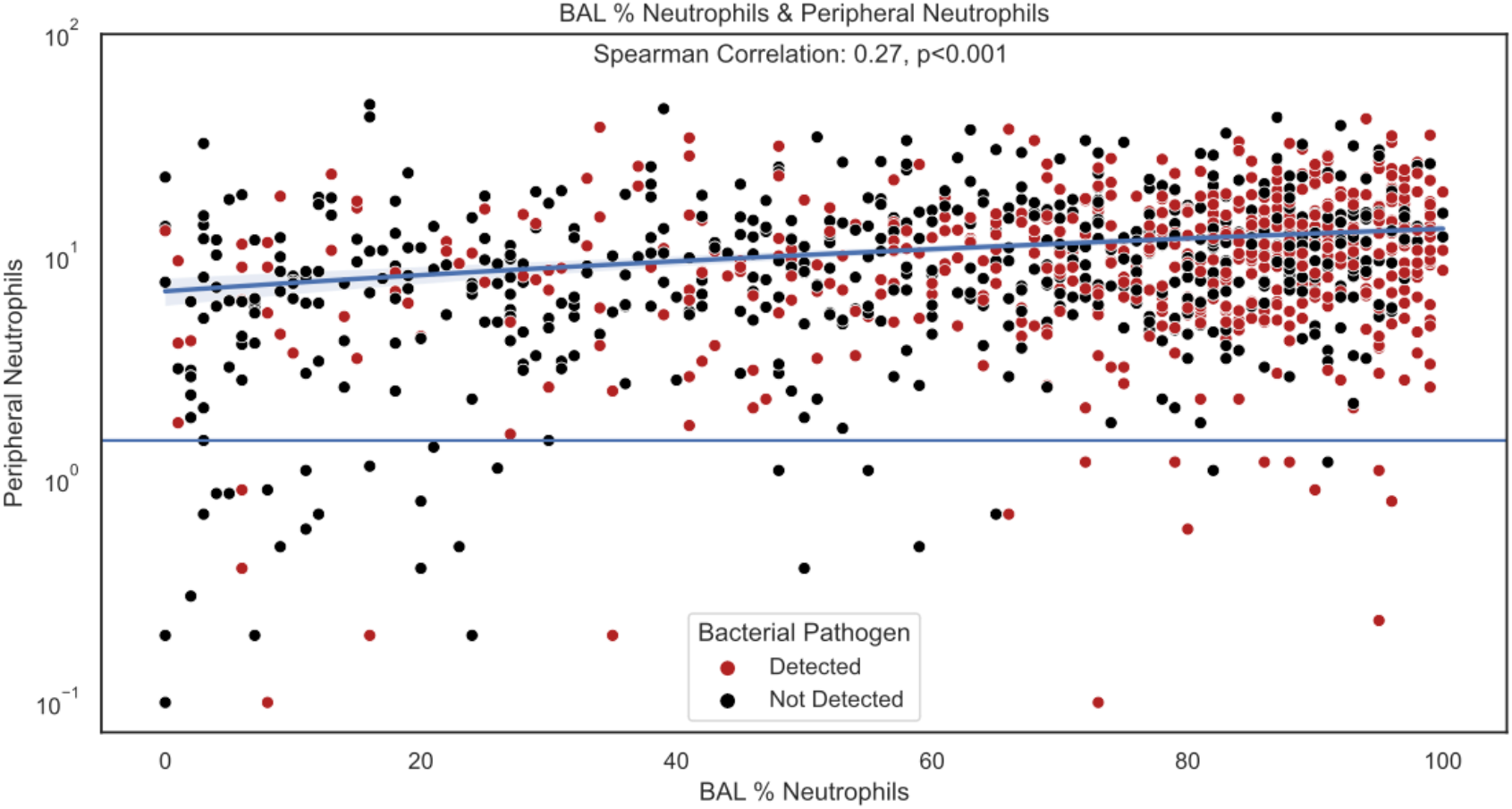
BALF % neutrophils across different peripheral blood neutrophil levels, immunocompromised hosts, and when sampling during neutropenia. BALF neutrophils and peripheral neutrophil count are correlated with Spearman Correlation of 0.27, p<0.001. Each dot represents a BAL sample, colored by whether a bacterial pathogen was detected (red) or not (black). The Y-axis is logarithmically scaled for ease of viewing the neutropenic BALs. A line at ANC = 1500 indicating our cutoff for neutropenia.

